# Sedentary time in adults with cystic fibrosis: A prospective observational cross-sectional study

**DOI:** 10.1101/2023.03.01.23286656

**Authors:** Ines Bishop, Robyn Cobb, Kathleen Hall, Suzanne Kuys

## Abstract

**Objective:** Adults with cystic fibrosis remain susceptible to comorbidities associated with high sedentary time, increasing their risk of poor health outcomes. Evidence about sedentary time in adults with cystic fibrosis is limited. This study investigated sedentary time and physical activity in adults with cystic fibrosis across disease severity groups and the relationship with clinical measures.

**Methods:** A SenseWear armband was worn by adults with cystic fibrosis, measuring sedentary time, time spent in moderate-vigorous physical activity and steps per day. Lung function, quadriceps strength, exercise capacity and health-related quality of life were assessed.

**Results:** On average, the armband was worn for 20 hours (Standard Deviation (SD) 6) over 6 (SD 1) days. Forty-eight participants (28 males) spent 815 (SD 379) minutes sedentary, 137 (SD 13) minutes engaged in moderate-vigorous physical activity and took 5660 (SD 2749) steps per day. Sedentary time did not vary across disease severity groups nor correlate with clinical measures.

**Conclusions:** Adults with cystic fibrosis spent a large proportion of time in sedentary behaviours, took insufficient steps, but conversely engaged in sufficient moderate-vigorous physical activity. Targeted interventions to break up sedentary time are required to reduce the risk of adverse health outcomes in this population.

**Impact:** Adults with cystic fibrosis spend a large proportion of their time sedentary, placing them at risk of future metabolic disorders. Interventions to break up sedentary time are needed.

## Introduction

Cystic fibrosis (CF) is an inherited multisystem progressive disease, with adult numbers now exceeding children.^1^ Improvements in early detection, and novel treatments, including management for the underlying pathophysiology of the CF transmembrane conductance regulator (CFTR) protein, have contributed to the survival rate, increasing to a median survival age of between 47.4 to 56.9 years in Australia.^2^ With increased life expectancy, people with CF become susceptible to comorbidities, including CF-related diabetes,^3^ osteoporosis,^4^ hypertension,^5^ and depression.^6^ Similarly, increased sedentary time has been associated with increased risk of all-cause mortality, cardiovascular mortality, cardiovascular disease, and type-2 diabetes.^7-10^ Evidence exploring sedentary time in adults with CF is limited. Investigating sedentary time in this cohort is essential to identify whether people with CF are at risk of developing these comorbidities. Additionally, the European position statement recommends including sedentary time, number of steps, and duration of physical activity for a comprehensive assessment of daily activity.^11^

In healthy adults, 150-300 minutes of moderate-vigorous physical activity (MVPA) weekly can reduce the risk of poor health outcomes.^12^ Previous studies investigating physical activity in the CF population focused on paediatric cohorts. These studies found that higher levels of physical activity were associated with a slower decline of lung function,^13^ greater muscle strength,^14^ improved quality of life and reduced risk of CF-related diabetes.^15^ Current studies in adults with CF have small sample sizes and focus on participants with mild and moderate disease but not severe disease.^16-18^ Assessing time spent in MVPA and number of steps along with sedentary time may provide insight into overall daily activity in adults with CF and potentially highlight areas of intervention for physiotherapy.

This study aimed to describe sedentary time, time spent in MVPA and number of steps taken per day in adults with CF across disease severity groups. In addition, this study aimed to investigate the relationship with clinical measures of lung function, quadriceps strength, exercise capacity, and health-related quality of life, including the direction and magnitude of this relationship. These findings may inform future management and delivery of physiotherapy services in adults with CF.

## Methods

### Design

A prospective observational cross-sectional study in adults with CF. The Prince Charles Hospital Human Research Ethics Committee (18908 HREC) approved this study.

### Participants

Inclusion criteria were adults (≥ 18 years) attending an adult CF outpatient clinic with confirmed diagnosis of CF. Participants across all disease severities were included and grouped based on disease severity classification; mild (>70% FEV_1_), moderate (40-70% FEV_1_), and severe (<40% FEV_1_).^19^ Adults with CF who were lung transplantation recipients, experiencing infective exacerbation requiring intravenous antibiotics, or presented with musculoskeletal or neurological conditions impacting physical activity participation were excluded from the study.

### Procedure

The attending physiotherapist at the CF outpatient clinic screened the clinic appointment list for eligible participants. Adults with CF meeting eligibility criteria were contacted via telephone, informed of the study and invited to participate. Participants attended their regular outpatient appointment at the adult CF clinic, where they were provided with an information sheet and consent form. Participants were notified that participation was voluntary and that they could withdraw at any stage without any consequences to their care. Once consent was granted, the treating physiotherapist assessed quadriceps strength and exercise capacity using the six-minute walk test (6MWT). Participants were fitted with the activity monitor and given the Cystic Fibrosis Questionnaire-Revised (CFQ-R) to assess health related quality of life to complete at home.

### Outcome measures

#### Primary outcome

The primary outcomes of sedentary time and physical activity were assessed using the SenseWear armband (SenseWear MF Bodymedia, Pittsburg, PA, USA). The SenseWear armband has been validated for measuring physical activity in this population (ICC 0.4, 95% CI 0-0.7).^16^ This non-invasive portable armband contains tri-axial accelerometers and sensors to measure galvanic skin response, heat flux, and skin temperature to estimate minute-by-minute energy expenditure^20^ and register upper arm movement during stepping^21^ to calculate the number of steps. The SenseWear armband was fitted to the participant’s upper arm to monitor physical activity. Participants were instructed to wear the armband for 24 hours a day over seven consecutive days, excluding water-based activities such as swimming or showering. All participants were instructed on the armband’s application, removal and use. Following seven days, the armband was posted to the clinic in a pre-paid envelope for data to be downloaded and analyzed using Bodymedia® software. Sedentary time, time spent in MVPA, and number of steps per day were extracted. Sedentary time, defined as time spent in a sitting or reclining position, was classified as < 1.5 metabolic equivalents (METS).^22^ Moderate-vigorous physical activity was classified as > 3 METS.^10^

#### Secondary outcomes

Secondary outcome measures included lung function, quadriceps strength, 6MWT and CFQ-R. Measures of lung function were recorded from the participant’s most recent spirometry test and expressed as forced expiratory volume in one second (FEV_1_) and percentage of predicted normal. Bilateral quadriceps strength was measured according to a standardized protocol, using a hand-held dynamometer stabilized by a strap attached to a set point on a raised plinth, with the participant’s knee in 70° flexion^2.3^ Resistance was applied to the anterior tibia, 5cm above the lateral malleolus and maintained for four seconds. Three trials for each leg were performed consecutively with 30 seconds to 1 minute rest between trials. The best measure from right and left quadriceps were recorded in kilograms to calculate average bilateral strength and strength corrected for body weight (best bilateral strength divided by body weight).

The 6MWT is a valid and reliable measure of exercise capacity in adults with CF^24^. The 6MWT was conducted according to a standardized protocol^25^ whereby participants were asked to walk as far as possible in six minutes along a marked flat straight corridor spanning 30 metres. Standardized instructions were delivered before the test, and encouragement was every 60 seconds for the duration of the test.^25^ Participants wore a pulse oximeter to monitor oxygen saturation and heart rate continuously. Participants rated their breathlessness from 0-10 using the modified Borg scale of breathlessness.^26^ Oxygen saturation, heart rate and breathlessness were recorded by the physiotherapist every minute for the duration of the test and one minute after completion. Results recorded included a six-minute walk distance (6MWD) (metres), highest score of breathlessness (/10), highest heart rate (bpm), lowest oxygen saturation (%) and use of supplemental oxygen.

The CFQ-R is a valid and reliable measure of health-related quality of life in people with CF.^27^ Participants answered 50-scaled questions exploring nine quality-of-life domains (physical functioning, vitality, emotional state, social limitations, role limitations/school performance, embarrassment, body image, eating disturbances, treatment constraints) and three symptom domains (weight, respiratory, and digestion). Questions were scored using 4-point Likert scales. Negatively phrased questions were recoded for scoring consistency, and scores were summed to generate a domain score then standardized. Results from each domain were converted to scaled scores ranging from 0-100, and a total percentage for each domain and the total score was calculated,^28^ with higher scores reflecting better quality of life.

### Sample size

A sample of 44 participants was required for this study. This was based on previous research,^16^ where a moderate correlation of 0.4 was observed between MVPA and FEV_1_. Using G Power 3.1.9.6, 44 participants were required for a two-tail test with a significance of 0.05 with 80% power. An additional 10% of participants were recruited to account for equipment malfunction and drop-outs.

### Statistical analysis

Statistical analysis was conducted using Statistical Product and Service Solutions (SPSS) version 26. All data were checked for normality using a Q-Q plot. Data were presented using descriptive statistics such as mean (standard deviation (SD)) or median (interquartile range). A one-way analysis of variance (ANOVA) was used to evaluate sedentary time, time in MVPA and number of steps per day across disease severity groups. Pearson’s correlation coefficient was used to investigate the correlation between sedentary time, time in MVPA and number of steps per day with lung function, quadriceps strength (bilateral strength and body weight corrected), 6MWD and CFQ-R score (total and each domain). Relationships were categorized based on strength with the following magnitudes: small <0.3, medium 0.3-0.5, large >0.5.^29^

## Results

### Participants

This study recruited 48 adults (28 males) with CF (Table 1). Data from five participants were not included in the analysis due to the inability to collect the activity monitor (*n* = 1), equipment malfunction resulting in data loss (*n* = 2), and poor participant compliance with wearing the armband (*n* = 2). Data from two participants were not included for quadriceps strength due to data loss. Average FEV_1_% predicted was 54% (SD 24). Fourteen participants (29%) were classified as having mild disease severity, 16 (33%) people had moderate disease, and 18 (38%) participants had severe disease. The average FEV_1_% predicted within disease severity groups was 86% (SD 7) for mild, 55% (SD 7) for moderate, and 29% (SD 5) for severe. The average quadriceps strength for right and left were 35.16 kg (SD 11.32) and 33.96 kg (SD 10.71), respectively.

**Table 1.**
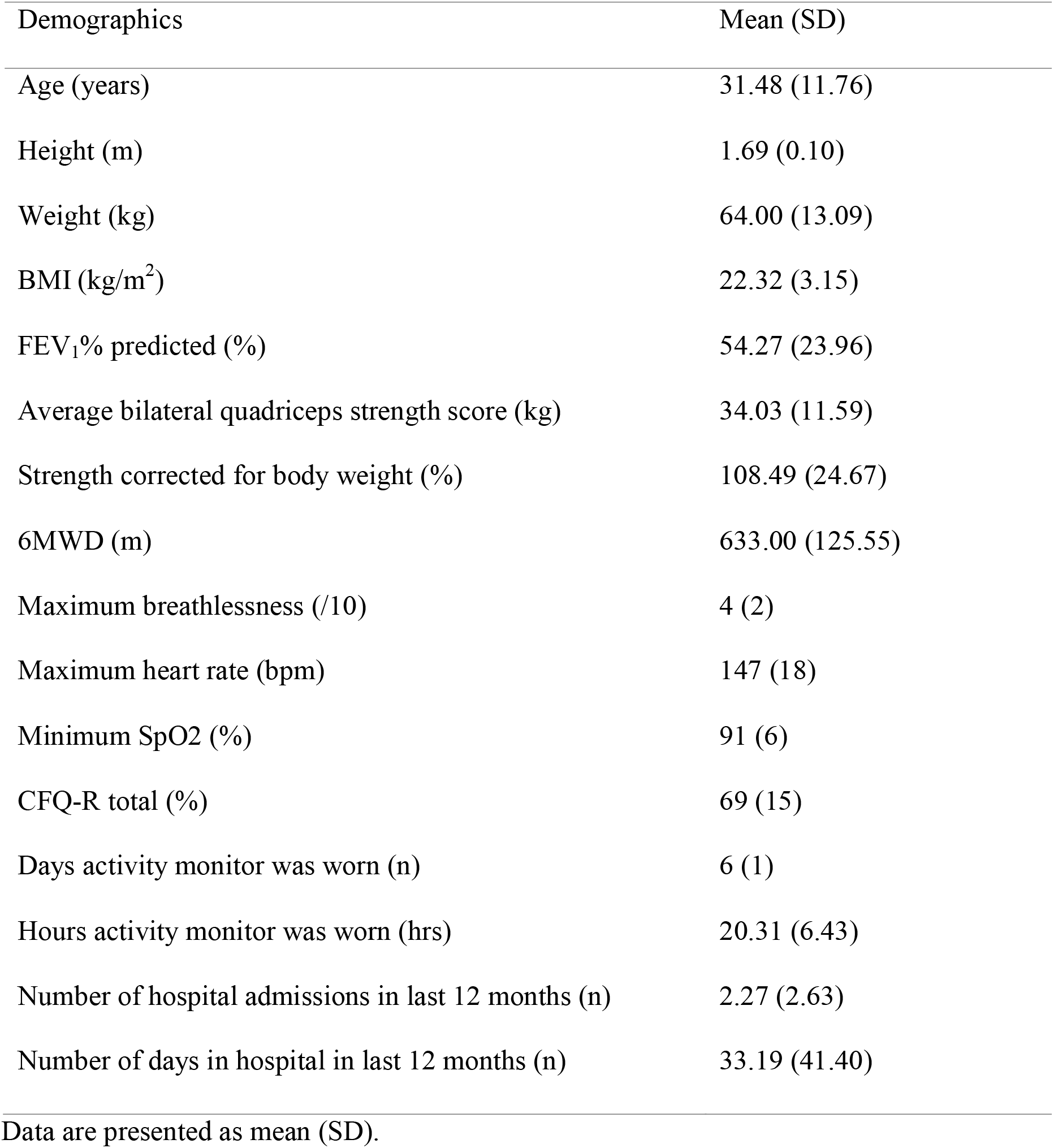
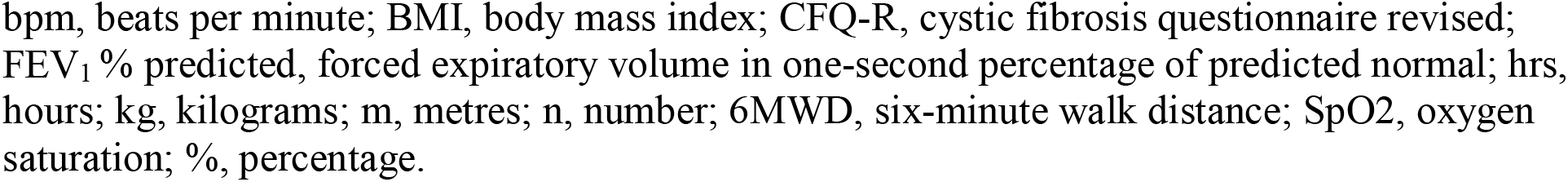
Participant Demographics

| Demographics | Mean (SD) |
| --- | --- |
| Age (years) | 31.48 (11.76) |
| Height (m) | 1.69 (0.10) |
| Weight (kg) | 64.00 (13.09) |
| BMI (kg/m<sup>2</sup>) | 22.32 (3.15) |
| FEV<sub>1</sub>% predicted (%) | 54.27 (23.96) |
| Average bilateral quadriceps strength score (kg) | 34.03 (11.59) |
| Strength corrected for body weight (%) | 108.49 (24.67) |
| 6MWD (m) | 633.00 (125.55) |
| Maximum breathlessness (/10) | 4 (2) |
| Maximum heart rate (bpm) | 147 (18) |
| Minimum SpO<sub>2</sub> (%) | 91 (6) |
| CFQ-R total (%) | 69 (15) |
| Days activity monitor was worn (n) | 6 (1) |
| Hours activity monitor was worn (hrs) | 20.31 (6.43) |
| Number of hospital admissions in last 12 months (n) | 2.27 (2.63) |
| Number of days in hospital in last 12 months (n) | 33.19 (41.40) |

Data are presented as mean (SD).
bpm, beats per minute; BMI, body mass index; CFQ-R, cystic fibrosis questionnaire revised; FEV<sub>1</sub> % predicted, forced expiratory volume in one-second percentage of predicted normal; hrs, hours; kg, kilograms; m, metres; n, number; 6MWD, six-minute walk distance; SpO<sub>2</sub>, oxygen saturation; %, percentage.

### Sedentary time and physical activity

Overall, the armband was worn for a mean of 6 days (range 3-7 days) for 20 hours (range 1-24 hours) per day. Table 2 presents sedentary time and physical activity across disease severity groups and the cohort as a whole. On average, participants spent 815 minutes (SD 379) being sedentary, 137 minutes (SD 130) engaged in MVPA and took 5660 steps (SD 2749) per day. Results from the ANOVA showed no statistical significance for sedentary time [F (31, 11) = 0.426, p = 0.970], MVPA [F (31, 11) = 0.925, p = 0.593], or number of steps [F (31,11) = 1.764, p = 0.160] across disease severity groups.

**Table 2.** Sedentary Time and Physical Activity Across Disease Severity Groups

| Physical Activity | Mild | Moderate | Severe | Total |
| --- | --- | --- | --- | --- |
| Sedentary time (mins) | 690.31<br>(369.88) | 908.57<br>(25.59) | 835.38<br>(422.87) | 815.35<br>(379.02) |
| MVPA time (mins) | 151.15<br>(60.30) | 129.00<br>(172.93) | 131.88<br>(134.12) | 136.77<br>(129.66) |
| Steps per day (n) | 7460 (2523) | 5649 (2974) | 4208 (1829) | 5660 (2749) |

Data are presented as mean (SD). mins, minutes; MVPA, moderate-vigorous physical activity; n, number.

### Relationship between sedentary time, physical activity and clinical measures

Table 3 reports the relationship between sedentary time, physical activity and steps per day with secondary measures of lung function, quadriceps strength, 6MWD and CFQ-R. Lung function had a large correlation with number of steps (*p* = 0.001) but was not significantly correlated with sedentary time (*p* = 0.445) or time spent in MVPA (*p* = 0.548). Average bilateral strength (*p* = 0.002) and strength corrected for bodyweight (*p* = 0.001) were significantly correlated with time spent in MVPA but not number of steps (*p* > 0.05) or sedentary time (*p* > 0.05). Six-minute walk distance was significantly correlated with time spent in MVPA (*p* = 0.024), number of steps (*p* = 0.013), but not sedentary time (*p* = 0.338). Correlations between physical activity and the CFQ-R domains were overall not significant except for with number of steps. Sedentary time was not correlated across any domains or with CFQ-R total (*r* = -0.088, *p* = 0.594). Time spent in moderate-vigorous physical activity was only significantly correlated with the eating domain (*r* = -0.351 *p* = 0.028). Number of steps was significantly correlated with the physical (*r* =,0.535 *p* = 0.001), role (*r* = 0.457, *p* = 0.005), emotional (*r* = 0.340, *p* = 0.034), social (*r* = 0.429, *p* = 0.006), and respiratory domains (*r* = 0.405, *p* = 0.011) as well as CFQ-R total (*r* = 0.431, *p* = 0.006).

**Table 3.** Pearson correlation coefficients (r) between sedentary time, physical activity and clinical measures

| Clinical Measures | Sedentary Time | MVPA | Steps Per Day |
| --- | --- | --- | --- |
| FEV<sub>1</sub>% pred | -0.120 (0.445) | 0.094 (0.548) | 0.505 (0.001) * |
| Bilateral quadriceps strength (kg) | 0.038 (0.811) | 0.476 (0.002) * | 0.178 (0.266) |
| QS% (kg) | -0.2456 (0.122) | 0.503 (0.001) * | 0.303 (0.054) |
| 6MWD (m) | -0.150 (0.338) | 0.344 (0.024) * | 0.375 (0.013) * |
| CFQ-R total (%) | -0.088 (5.94) | 0.039 (0.815) | 0.431 (0.006) * |

\*p < .05
Data are presented as correlation coefficient (p-value).
CFQ-R, cystic fibrosis questionnaire revised; FEV<sub>1</sub>% pred, forced expiratory volume in one-second percentage of predicted normal; kg, kilograms; m, metres; QS%, quadriceps strength corrected for bodyweight; 6MWD, six-minute walk distance; %, percentage.

## Discussion

This study showed that adults with CF engaged in high amounts of sedentary time, spent a considerable amount of time in MVPA, and took low numbers of steps per day. There was some variation across disease severity groups, though this was not statistically significant. Sedentary time was not related to any clinical measures. However, both time spent in MVPA and number of steps taken per day positively correlated with quadriceps strength and exercise capacity.

This high amount of sedentary time is concerning. On average, participants in this study spent approximately 13 hours per day sedentary. Sedentary time is defined as time spent lying or sitting, excluding sleeping time^22^ If sleeping time is accounted for, participants in this study spent little time doing anything else apart from being sedentary or sleeping. Increased sedentary time has been associated with increased risk of cardiovascular disease, type-2 diabetes^7^, and more than ten hours of sedentary time has been associated with a 1.5 times higher risk of death.^8^ Therefore, adults with CF who are spending large amounts of time sedentary are potentially increasing their risk of developing other chronic diseases and increased mortality. With the introduction of Transmembrane Regulator modulator therapies over the last decade, the risks of weight gain^30^ and metabolic complications such as hypertension^31^ in adults with CF are already being documented as complications to be managed. Additionally, obesity is a known risk with co-existing sedentary behaviour. Physiotherapists can utilize education to emphasize breaking up sedentary time with light physical activity, as the World Health Organization^32^ recommends, to reduce the risk of metabolic complications. Future delivery of physiotherapy services should include interventions and education as an adjunct to transmembrane regulator modulator therapies.

Adults with CF in this study spent over 2 hours engaged in MVPA daily (> 3,0 METs). This suggests that participants more than achieved the recommended physical activity guidelines for time spent in MVPA.^32^ Additionally, this finding is more than the 31 minutes of daily MVPA previously reported in adults with CF.^16^ However, this data should be interpreted cautiously due to different classifications of MVPA used. It has been suggested that adults with CF have higher resting energy requirements and therefore experience higher energy expenditure with physical activity compared to healthy populations for the same task.^33^ Classification of MVPA has not been standardized within the cystic fibrosis community, but a higher threshold of >4.8 METS for moderate intensity has been identified as more appropriate by Ward et al. 2013,^34^ Cox et al. 2016 ^16^ and Troosters et al. 2009.^18^

In keeping with the potential for overestimation of physical activity, adults with CF in this study took a low number of steps per day, on average around 5600. This is well below the recommended target of 10,000 steps per day^35^ compared to previous studies, which recorded more than 9000 steps daily^17,18^ The discrepancy between these studies requires exploring. Previous studies had smaller sample sizes^18^ and did not include adults with severe disease,^17^ which might account for the differences. However, participants with mild disease in the current study did not meet these targets either. It appears this group of adults with CF are not taking sufficient steps per day for health benefits, which should be addressed through education by health professionals managing this population.

Although not statistically significant, there appears to be some relationship between sedentary time, time spent in MVPA, number of steps per day and disease severity groups. Sedentary time was high regardless of disease severity, more than 10 hours on average. Unsurprisingly, adults with mild disease appeared to spend the least time being sedentary, also spending more time in MVPA and taking more steps per day compared to adults with moderate and severe disease. Interestingly, the severe disease group spent the largest duration of time being sedentary, took the least steps but still achieved the guidelines for time spent in MVPA. A possible reason for this could be that adults with severe disease have a higher energy expenditure with physical activity for everyday tasks and therefore appear to spend more time in MVPA when they are performing light activities only.^18^ The CF standards of care and clinical practice guidelines recommend 30-60 minutes of MVPA per day as aerobic exercise.^36^ Exercise can enhance sputum clearance^37^ and is recommended to be used in conjunction with other airway clearance techniques.^38^ In this adult centre, adults with severe disease are encouraged to engage in regular exercise as an adjunct to airway clearance. However, fatigue from exercise at MVPA may cause patients to spend the remainder of time sedentary. This study showed that sedentary time was not related to any clinical measures. This suggests that the finding of high sedentary time needs to be addressed in all adults with CF regardless of disease severity or clinical performance.

A strength of this study is the replicability. The clinical measures assessed are routinely conducted in people with CF.^38^ The SenseWear armband was easy for physiotherapists and participants to use. Therefore, using these clinical measures and the armband could be repeated in other clinical settings. This study used an objective measure of sedentary time and physical activity, reducing the risk of overestimation as seen with subjective measures.^17, 39^ Since sedentary time and physical activity were assessed in the outpatient setting over 5-7 days and across all disease severity groups, overall findings were reflective of habitual daily activity and accounted for the variability of activity across days and disease severity groups.

### Limitations

One limitation is that this was a single-centre study. However, this study had a similar distribution of participants across genders and disease severity groups and was conducted in the sole state-wide centre for adults with CF in Queensland, making it reflective of CF cohorts in Australia.^2^ Limitations associated with the SenseWear armband, including that it only measured land-based activities and was sometimes removed for social reasons, may have led to an underestimation of overall physical activity in some participants.

## Conclusion

This study demonstrated that while adults with CF spend a large portion of the day in sedentary behaviour and perform fewer steps per day than recommended, they conversely participate in sufficient MVPA. The high amount of sedentary time potentially increases the risk of all-cause mortality in adults with CF. This study highlights the need for increased education and prevention of sedentary time in this population to promote healthy behaviours. This is of increasing importance with the introduction of CFTR modulator therapies which play a significant role in correcting CFTR dysfunction leading to the amelioration of weight loss.^30^ The role of physical therapy within this population should target education on the health risks associated with increased sedentary time and provide strategies to mitigate these, such as breaking up sedentary time and replacing it with light physical activity. Future studies investigating interventions to break up sedentary time in adults with CF are warranted to assess the duration and frequency of breaks to promote positive health outcomes.

## Data Availability

All data produced in the present work are contained in the manuscript.

## Acknowledgements

The authors wish to acknowledge the staff of The Prince Charles Hospital and Australian Catholic University involved in this study.

